# Perceived Impacts of Generational Knowledge on Female Genital Mutilation and Barriers to Awareness Programs in Ibadan, Oyo State

**DOI:** 10.64898/2026.09.03.26362187

**Authors:** Semiyu Gbadebo Hammed, Enemona Jacob

## Abstract

**Introduction:** Nigeria is still one of the countries in the world that practices Female Genital Mutilation (FGM), which remains a significant public health issue in Nigeria, particularly in Ibadan, with a prevalence rate of 38%. Despite increased awareness and advocacy, FGM persists as a rite of passage, chastity, or cultural custom. In this study, participants revealed generational knowledge gaps of FGM and barriers to the effectiveness of awareness programs.

**Methods:** This was a narrative inquiry study conducted between August 2024 and December 2024 among female young adults of reproductive age (18-49 years) living across different local government areas of Ibadan metropolis. A purposive and snowballing sampling technique was adopted, where semi-structured in-depth interviews were used. Interviews were audio recorded and transcribed verbatim, where insights into participants’ experiences, perspectives, and attitudes impacted by awareness programs, as well as the interaction of cultural phenomena with public health education, were discussed and analysed through thematic analysis to present findings.

**Results:** The participants were able to disclose that awareness programs in schools and community-based initiatives have proven to empower younger generations, but bridging generational knowledge gaps remains a challenge, with barriers like entrenched cultural norms, limited geographical accessibility, insufficient funding, and weak enforcement of anti-FGM laws, and suggestions on how awareness programs against FGM can be more effective.

**Conclusions:** Awareness programs on younger generations are effective in tackling the prevalence of FGM, although they still face some entrenched cultural norms. The integration and sustenance of culturally sensitive messages into the education and healthcare sectors, both in rural and urban areas, will be of great help to counter the entrenched cultural norms of people still practicing them. Similarly, the implementation and enforcement of laws and policies by the government, and the proper execution of sanctions against the practice of FGM in society, should be adopted.

## Introduction

Female genital mutilation (FGM), a traditional practice among women in close-knit communities, faced resistance from European missionaries and colonizers during the colonial era, who viewed it as barbaric and incompatible with Western values.^53 60 86^

In the modern era, the global community has taken a strong stance against FGM, particularly after the adoption of the Universal Declaration of Human Rights in 1948, which recognized the rights of women and girls.^36 46 126^ The World Health Organization (WHO) defined FGM as a violation of human rights, emphasizing its harmful consequences for the physical and psychological well-being of girls and women.^36 44^ In regions such as the Horn of Africa, including Somalia, Ethiopia, and Eritrea, FGM has been a long-standing tradition.^19 34 137^ Similarly, in countries like Egypt, Sudan, and Mali, the practice remains prevalent, despite increasing awareness and efforts to end it.^5 9 52^

In some West African cultures, FGM is associated with religious beliefs, particularly in communities where Islam is predominant, although the practice is not condoned by Islamic teachings.^24 66 119^ In other cases, FGM is closely linked to local beliefs about purity, cleanliness, and the preservation of family honor.^106 130^ The procedure is often performed by traditional practitioners or midwives, and in some regions, it is carried out in unsafe, unsanitary conditions, leading to significant health risks such as infection, hemorrhage, and complications during childbirth.^8 51 113^

West African countries have made significant strides in addressing FGM. International organizations such as UNICEF, the WHO, and the United Nations have worked in collaboration with local governments and non-governmental organizations (NGOs) to raise awareness, provide education, and pass laws to prohibit FGM.^71 95 101^ However, the practice persists in many rural and remote areas, where it continues to be seen as a vital part of cultural heritage and societal norms.^6 107^ In Nigeria, FGM is often performed during childhood, typically between the ages of 5 and 14.^94 97 98^ Nigeria is working to eradicate FGM through campaigns and advocacy by local and international organizations, as well as religious and community leaders,^11 63^ but the practice remains widespread, particularly in rural areas where traditional beliefs are deeply ingrained.^2 12 106^

Oyo State, located in southwestern Nigeria, is home to the Yoruba people, one of the largest ethnic groups in Nigeria.^132^ The practice of FGM in Oyo State, like in many other parts of Nigeria, is deeply embedded in cultural and traditional practices. In the Yoruba culture, FGM has been practiced for centuries as a way to preserve virginity, control women’s sexuality, and ensure a successful marriage.^4 102^ It is often seen as an important rite of passage that marks a girl’s transition into womanhood. However, Oyo State is leading efforts to combat FGM in southwestern Nigeria, with NGOs, health organizations, and the Nigerian government promoting alternative rites of passage.^3 62 103^

In urban areas like Ibadan, the state capital, awareness of the dangers of FGM has led to a decrease in its prevalence.^105 129^ However, rural communities in Oyo State face challenges in eliminating FGM due to cultural pressure,^69^ and the government collaborates with organizations to enforce legislation, educate, and support community-led efforts.^69 101^

The practice of female genital mutilation has been the subject of numerous research, some of which concentrated primarily on the prevalence of this practice, in which some studies looked at attitudes towards female genital mutilation across generations, and their method was quantitative, and their data were retrospective, therefore only associations could be made, and participants’ actual experiences could not be qualitatively ascertained.^55 56^ Consequently, there is a lack of comprehensive understanding regarding the effectiveness of awareness programs in reaching and engaging this demographic group.^30 77 78 136^

An FGM study situation analysis conducted by UNFPA^133^ in Nigeria has demonstrated that, in order to raise awareness among pregnant women in various states, primary healthcare centers have integrated FGM education classes into prenatal and postnatal services at the community-based healthcare level. Despite the inclusion of FGM-related courses in the curricula of health training institutions,^39^ ^45 37^ only a small percentage of medical professionals have been impacted, suggesting a knowledge gap in Nigeria’s FGM prevention and management.^100 45^ Nonetheless, substantial gaps remain in comprehending the continued impact of awareness programs on generations and the barriers to the effectiveness of awareness programs of this practice despite heightened awareness among teenagers and young adults (18-49 years) in Ibadan. This study’s relevance transcends the local context of Ibadan, with the capacity to inform and impact policy and practice at state, national, and global levels, and will provide suggestions for enhancing future intervention strategies by clarifying the effectiveness of different methods used in awareness efforts via data-driven and evidence-based results.

## MATERIAL AND METHODS

### Research Theory

To support this study, the socio-ecological model (SEM) originally proposed by Bronfenbrenner in 1979 was adopted.^114^ The SEM offers a comprehensive framework for understanding the multiple influences on FGM practices and the effects of awareness programs in Ibadan, Nigeria. In the context of FGM, the SEM provides a detailed approach to examining how awareness campaigns influence young women’s knowledge, attitudes, and behaviours across various levels, such as individual, interpersonal, community, organizational, and policy levels.^114^ This approach enables the design of interventions that can create synergies between individual and social-environmental factors (as described in the SEM), potentially leading to more effective and sustainable behaviour change strategies in the fight against FGM.^135^

### Research Philosophy

This study uses interpretivism and social constructivism to explore how young women in Ibadan, Nigeria, perceive female genital mutilation (FGM) awareness programs. The research acknowledges that participants’ perceptions and experiences may vary based on their cultural background, education, and exposure to awareness initiatives. This approach allows for a deep exploration of how awareness programs interact with social norms and cultural practices, providing contextually rich and methodologically sound insights.

### Researcher characteristics and reflexivity

Researchers’ personality characteristics may have impacted this study’s design, conduct, and interpretation. I’m a male researcher who used to reside in Ibadan, Oyo State, the study community. This previous stay offered cultural familiarity, contextual awareness, and understanding of the language, which made it easier to access participants and promoted rapport-building on sensitive topics like female genital mutilation (FGM). The insider status did, however, also carry the risk of presumptive shared understandings, which was deliberately worked to challenge through reflexive practice.

Given the gendered character of FGM, participant interactions may have been influenced by my gender. Some female participants might have given answers that were cautiously phrased or with little candour. In order to address this, the researcher conducted interviews in a courteous, nonjudgmental manner and, where necessary, let participants lead the conversation.

### Study Setting

This study was conducted in Ibadan, Oyo State, Nigeria. Ibadan, the capital and most populous city of Oyo State in Nigeria, is the country’s third-largest city, following Lagos and Kano. With a population of 2,649,000 as of 2021^7^, it is one of the country’s largest cities in terms of geographical area. Ibadan is located in southwest Nigeria, 128 kilometers northeast of Lagos and 530 kilometers southwest of Abuja. It acts as a link between countries’ coastal regions and hinterlands.^7 16^

The principal inhabitants of the city are the Yoruba people, and various communities from other parts of the country (Ibadan, 2024). There are 11 local governments in the Ibadan metropolitan area, consisting of five urban local governments in the city and six semi-urban local governments.

### Study Design

This study uses narrative inquiry as a qualitative research design to explore the experiences of the participants with FGM and related awareness programs. Narrative inquiry prioritizes perspectives of the participants and allows for a deeper understanding of cultural norms and awareness campaigns.^32 74^ It focuses on personal stories, addressing the complex interplay of rights, gender, and health in the context of FGM. This method ensures methodological rigor and transparency while maintaining ethical standards.

### Study Participants: Inclusion/exclusion criteria

This study focused on young and older women, primarily from Ibadan, Nigeria, between the ages of 18 and 49, who had either participated in awareness campaigns regarding female genital mutilation (FGM) or had undergone the procedure. The population was chosen because it is both a target group for FGM interventions and a possible agent of community change. Participants who did not meet all these criteria were excluded from participating in the study.

### Participants Recruitment

A variety of online platforms, including digital channels like Telegram and WhatsApp, were used to recruit participants. In order to facilitate a formal and ethically acceptable recruitment process, participants were given instructional posters and invitation messages, and digital consent was signed before the scheduling of interviews through Microsoft Teams. Participants obtained a consent form and participant information sheet via Qualtrics, which included information on the study’s objectives, voluntary participation, confidentiality guarantee, and withdrawal protocols.

The study location, Ibadan, Oyo State, is a Nigerian state with a high level of educated individuals. This indicates that the target audience, which is between the ages of 18 and 49, is a highly educated generation that has grown up with technology,^50^ and the literacy rate is very high. They are a mix of Gen Z and the millennial generation, both of which are recognised for their education and familiarity with digital media.^50^ Since the participants are from the Y and Z generations and Oyo State is known for having a high level of education, they were judged to be educated and so qualified to take part in the study’s online interview.

### Study Population and Sampling

The study used purposive and snowball selection techniques to recruit participants relevant to its objectives, with a focus on women with FGM-related experiences.^21 25 89^ Snowball sampling expands the scope while retaining relevance by enabling more participants through recommendations.^111^

The sample size of 22 participants out of the 44 registered people who showed interest in participating in the study interview was between the ages of 18 and 49 years (this was led by data saturation). In thematic and narrative research, data saturation, also known as “information redundancy,” is a commonly recognised cutoff point for figuring out an adequate sample size.^40^ Participants were selected from 7 local governments in Ibadan and educated to university level, which helped to assure thoroughness while balancing methodological thoroughness with practical limits.^128^ ^138^ Thirteen of them were between the ages of 18 and 35, nine were between the ages of 36 and 49, five were students, nine worked for themselves, and eight were employed in other industries.

### Ethical approval

The study received approval from the University of Hertfordshire Health, Science, Engineering, and Technology Ethics Committee (Protocol number: cLMS/PGT/UH/05727; dated August 12, 2024), in accordance with established protocols. Participants were informed and gave their consent through a secure online platform (Qualtrics), where their personal information was replaced with anonymous codes. Participants were informed of their voluntary participation and their right to withdraw. We set up data encryption and secure storage mechanisms, and we used anonymous IDs to keep participants’ privacy safe. The processes were finished during the ethical application phase, which came before getting clearance from the ethics commission after a thorough review.

### Data Collection Procedure

This study used online semi-structured interviews to gather data on the complex |making it suitable for exploring sensitive topics like FGM. The semi-structured format allows for guided conversations and allows participants to elaborate on their perspectives, allowing for new themes and insights. Online interviews reduce logistical barriers, increase accessibility, and maintain anonymity, which is crucial when discussing culturally sensitive practices. The interview instrument was optimized for clarity and comprehension, minimizing ambiguity and addressing cultural and emotional sensitivities. This approach ensured that the voices of the participants were at the forefront of the research, effectively capturing the relationship between cultural norms, personal experiences, and awareness programs’ impact.

### Data collection instruments

The interview guide utilised in this research was specifically created for this study and is included in the supplemental materials. The interview guide was crafted to guarantee clarity and cultural sensitivity while fostering a secure environment for participants to discuss their experiences. The interview was conducted in English, and participants’ responses were recorded. Sample questions for the interview include: “What are the main reasons for FGM practices in your community?” “What role do you think awareness programs can play in the prevalence of FGM?” “How do you feel about awareness programs having any impact on the practice of FGM in your community?” and “Have you witnessed changes in attitudes due to awareness programs?” On a scale of 1-10, can you rate the impact of the FGM awareness program on your community?”

“What is now your opinion about the practice of FGM that you may likely pass on to upcoming generations?” These questions allowed for an in-depth understanding and data collection about awareness programs. The interviews were held via Microsoft Teams, allowing for personal time convenience and a duration of 45-60 minutes. The study aimed to ensure responsible data storage and management of personal data and recordings, upholding participant confidentiality and trust. Data was securely stored on password-protected electronic devices or servers, with restricted access to others. Data backups were performed to prevent losses in case of hardware failure or other unforeseen circumstances. At the end of the retention period, personal data and recordings were securely destroyed following data protection regulations and institutional policies.

Participants were informed about these confidentiality measures in the Participant Information Sheet before obtaining consent.

### Unit of Study

Participants were selected across 7 local governments and educated to university level, 13 of them were between 18-35 years, while 9 were between 36-49 years; 5 students and 9 were self-employed, while 8 were employed in different sectors, 5 were singles, and 17 were married.

### Data Management

The study complied with ethical and regulatory standards while ensuring effective data management and storage methods.^116^ Personal information and recordings were retained solely for research purposes. Upon the verbatim transcription of each, as advised by qualitative research,^47^ they were securely destroyed, and physical copies were shredded, ensuring compliance with the General Data Protection Regulation (GDPR) and institutional policies.^70^

### Data Analysis

Thematic analysis was chosen as the method for the study due to its flexibility and applicability to exploring complex, context-specific phenomena, such as FGM and awareness programs. This approach enables the identification of patterns and themes within qualitative data, providing a systematic framework for interpreting perspectives of the participants.^29^ This method facilitates a thorough analysis of cultures, individuals, and groups, serving as an effective tool for comprehending society’s relationship with its narratives.^109 112^

The study adhered to Braun and Clarke’s six-phase process for data analysis: (1) familiarisation with data, (2) generating initial codes, (3) searching for themes, (4) reviewing themes, (5) defining and naming themes, and (6) writing the report.^26 27^ The initial step entailed familiarising oneself with the data by examining transcripts from Microsoft Teams and storing them in the secure OneDrive of the University of Hertfordshire. This stage enables the researcher to thoroughly examine the data and acquire unbiased preliminary insights into the content.^48 61^ The subsequent phase entailed coding the data and systematically labelling associated concepts and patterns.^92 123^ The researcher employed manual coding to establish a personal connection with the data and enhance the recognition of nuanced insights. The codes were aggregated into clusters to establish initial themes, emphasising participants’ attitudes towards FGM and their connections to effective awareness campaigns.^28 139^ The themes underwent iterative review to guarantee coherence and relevance to the research questions. The process entailed verifying the alignment of themes with the coded data and refining them to resolve any overlaps or inconsistencies. Themes were defined and named according to their scope and significance, ensuring accurate conveyance of key insights from the data. The concluding step entailed synthesising the themes into a unified narrative. This phase involved the incorporation of direct participant quotes to exemplify the themes and validate the findings through data grounding. The report was organised to meet the research objectives, connecting themes to the wider cultural and social context of FGM.

### Study Trustworthiness

The study followed rigorous standards for validity and reliability, adhering to Lincoln and Guba’s (1985) trustworthiness criteria. Four key components were incorporated: credibility, transferability, dependability, and confirmability. Triangulation confirmed the findings’ credibility through multiple data sources while member-checking for consistency in themes and patterns. Results were presented with direct quotes, highlighting participant perspectives. Comparing responses across participants further confirmed the reliability and depth of the findings.

This study ensured transferability by providing transparent descriptions of its context, methodology, and participant characteristics. It also established dependability through meticulous documentation of the research process, maintaining an audit trail for external review. Confirmability was supported by a reflexive approach, acknowledging personal biases and ensuring interpretations were grounded in participant data, minimizing subjectivity and safeguarding the objectivity of the research outcomes.

The study adhered to the Consolidated Criteria for Reporting Qualitative Research (COREQ) guidelines to improve its trustworthiness. These guidelines ensure transparent disclosure of methodological decisions and ethical considerations. The study aims to produce credible, transferable, and reliable findings that contribute to understanding FGM and awareness programs’ effectiveness.

## Results

### Findings

A total of 44 participants, aged between 18-49 years, who showed interest in participating in the study were invited from all local government areas in the Ibadan metropolis to participate in a Microsoft Teams interview for the study. Out of these, 22 participants (50%) subsequently took part in the interview, made up of 12 graduate students and 10 post-graduate students. Recruitment was place in seven local government (LG) regions in the Ibadan metropolis: Akinyele LG, Egebda LG, Ido LG, Ibadan SW LG, Ibadan SE LG, Oluyole South LG, and Ona Ara LG.

The analysis of this study was carried out through thematic analysis, following Braun and Clarke’s framework, and was used to identify and organize two central themes, which are Knowledge Transfer to Younger Generations and Barriers to the Effectiveness of Awareness Programs. From these themes, sub-themes were also derived from participants’ interviews, aligning with the study objectives. This process involved coding participants’ responses and grouping related ideas into themes.

**Note:** We used the SRQR reporting guideline to draft this manuscript, and the SRQR reporting checklist when editing, included in supplement A’

**Table 1:**
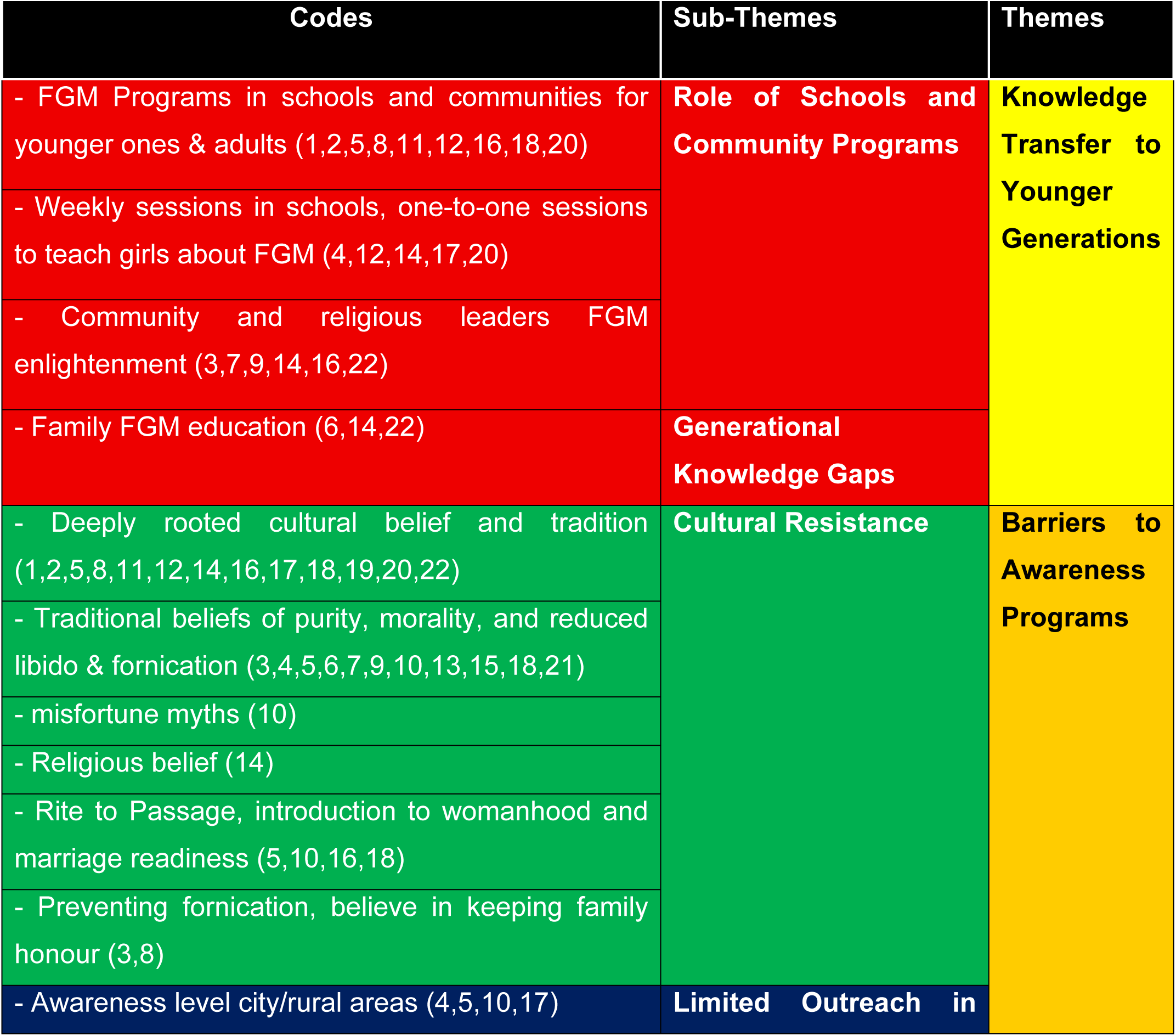
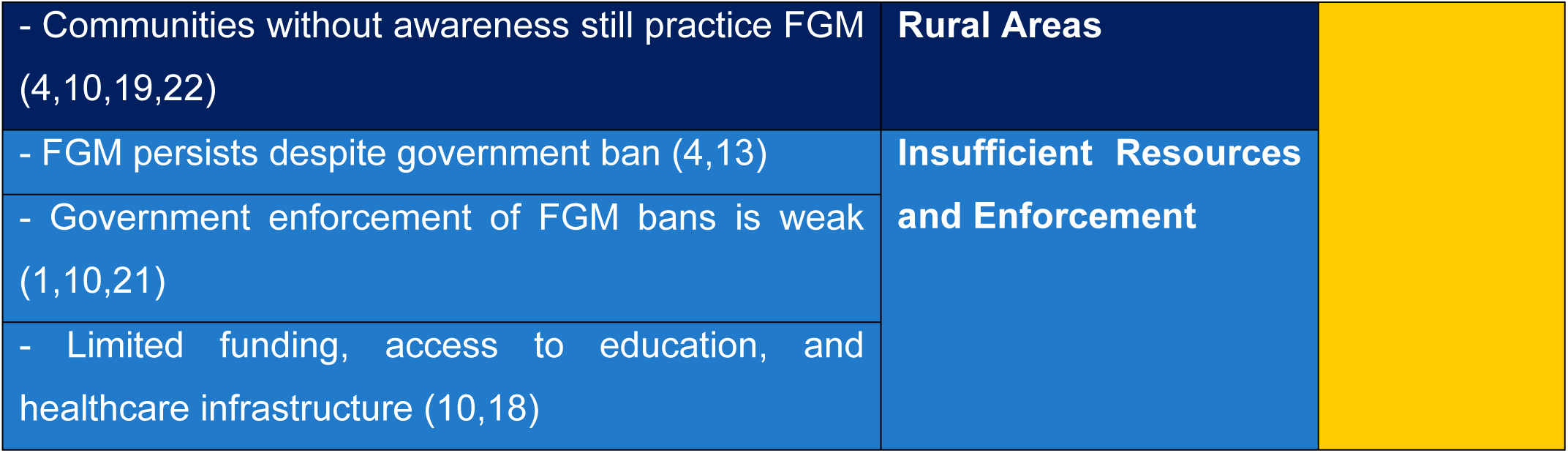
The codes and themes generated from participants’ responses are shown in the table below.

### Theme 1. Knowledge Transfer to Younger Generations

The transfer of knowledge across generations is critical to the long-term eradication of FGM. Awareness programs targeting young people, particularly in schools and community centers, have emerged as powerful tools for disrupting the intergenerational transmission of harmful practices. However, the study also highlights gaps in generational knowledge transfer, as older family members often hold traditional views that conflict with the anti-FGM messages being imparted to younger individuals. This theme addresses how awareness programs influence the continuity or cessation of FGM practices across generations.

### Subtheme 1: Role of Schools and Community Programs

Schools and community-based programs are instrumental in equipping younger generations with knowledge about FGM, empowering them to make informed decisions and challenge harmful traditions. These programs provide safe spaces where adolescents can learn about the health risks, psychological consequences, and legal ramifications of FGM, often engaging participants through interactive discussions and relatable examples.

Participants highlighted the effectiveness of these initiatives in urban areas, where access to schools and public campaigns is more common.

> “Programs in schools and communities help spread the message against FGM,”—p16

emphasizing the role of formal education in fostering critical thinking about FGM.

> “We used to have different placards back then in school showing dangers of body mutilation, especially that of facial and private parts for females. So I can never do such to my daughter, and I will never encourage such”—p7

Similarly, another participant noted;

> “Awareness has helped reduce the practice. It’s more common in cities than rural areas.” —p5

These insights show that urban settings provide an advantage in delivering consistent anti-FGM messaging.

Despite awareness campaigns, participants feel that more robust educational programs are necessary to fully address the issue.

> “Awareness is not enough, we need more education.”—p1
>
> “I don’t think I have ever heard any jingles with regards to that on the radio”— p11
>
> “I am not sure if schools still have educational placards or banners like we used to have in the early 90s. These are what will put a stop to these things”—p7

Expanding school-based and community initiatives to underserved regions is crucial for ensuring that all adolescents receive the same opportunities to learn about FGM, ultimately promoting a generational shift in attitudes toward the practice.

### Subtheme 2: Generational Knowledge Gaps

While youth-focused awareness programs have successfully influenced younger generations, bridging generational gaps remains a significant challenge. Older family members, particularly mothers and grandmothers, often uphold FGM as a cultural obligation, complicating the transfer of anti-FGM knowledge within households. This generational divide highlights the need for multi-generational education approaches. Another participant emphasized the importance of involving entire families in education, stating:

> “Educating families together reduces resistance.”—p14

This approach ensures that older family members, who are typically decision-makers, also understand the health risks and human rights violations associated with FGM. Without their buy-in, younger individuals may face pressure to conform despite their awareness of the practice’s consequences. A participant remarked,

> “Communities without much exposure still practice FGM,”—p22 reflecting how isolation and limited access to information perpetuate traditional beliefs among elders.
>
> “As it is, my mother never told me any valid reasons for doing that to me, all I know is a family tradition. Thank God my husband’s family doesn’t practice such a wicked tradition”—p13
>
> “Engaging community leaders has helped reduce FGM practices.”—p9
>
> Addressing generational knowledge gaps requires targeted outreach and inclusive education efforts that involve both adolescents and their family members in shared learning environments, promoting collective shifts in perspective.

### Theme 2. Barriers to the Effectiveness of Awareness Programs

Despite their success, awareness programs face significant barriers that limit their reach and impact. These barriers include entrenched cultural norms, limited geographical accessibility, insufficient funding, and weak enforcement of anti-FGM laws. Cultural resistance, in particular, remains a dominant obstacle, as FGM is deeply rooted in social and religious traditions. The lack of adequate resources and outreach in rural areas further exacerbates the problem, as these communities often have limited exposure to anti-FGM initiatives. This theme encapsulates the challenges that awareness programs encounter in reaching diverse populations and achieving sustainable change.

### Subtheme 1: Cultural Resistance

Cultural beliefs remain one of the most significant barriers to the effectiveness of awareness programs. Many participants described how FGM is viewed as a rite of passage, a means of preserving purity, and a method of controlling female sexuality, making it difficult for individuals to reject the practice.

> “I know is a family tradition. Thank God my husband’s family doesn’t practice such a wicked tradition”—p13
>
> “It’s done to control women and prevent sexual urges. So I had that it doesn”t make one to be promiscuous…it that through, I don’t think so, I feel normal sexual urge as a lady. I was never a virgin when I get married despite being mutilated.”—p19
>
> Traditional beliefs hold that FGM keeps girls pure.”—p9

These insights highlight the challenge of challenging deeply rooted cultural norms that view FGM as essential.

### Subtheme 2: Limited Outreach in Rural Areas

Participants pointed out that awareness programs often fail to reach rural communities, where FGM prevalence remains high due to isolation and lack of access to education or healthcare services.

> “I don’t think anyone in the city here will do that, I was born in village and I know that’s the reason they did it for me any sister then, I don’t think any of us can do that to our daughters because of our exposure”—p17
>
> “My grandma still believes in the practice o, I remember when I travel home for visit she joking asked if I had done FGM on my daughter. I was like oh my God, in this century”—p10
>
> “Despite the government ban, FGM still occurs in some communities.”—p13
>
> “Awareness programs are more common in cities than rural areas.”—p22

This theme reflects the geographic disparities in access to anti-FGM initiatives and underscores the importance of expanding outreach efforts.

### Subtheme 3: Insufficient Resources and Enforcement

Participants highlighted inadequate funding, resource limitations, and weak enforcement of anti-FGM laws as significant barriers to the success of awareness programs.

> “People still do it because they think no one will stop them.”—p5
>
> “Government bans exist, but enforcement is weak.”—p13
>
> “We hear of laws, but we hardly see any action taken. If someone is caught, they are warned and released.”—p7
>
> “There is no follow-up or monitoring. After the campaigns, everything goes quiet.”—p9
>
> “Even the police do not take FGM cases seriously. They say it’s a family matter.”—p11
>
> “We need resources—vehicles, staff, materials—to reach more communities. Right now, we do what we can with little.”—p4
>
> “Many of us want to help stop it, but there is no financial support or incentive to keep pushing.”—p16

This finding emphasizes the need for stronger legal frameworks and resource allocation for anti-FGM campaigns, highlighting the demotivation of community volunteers and stakeholders due to insufficient funding. It also highlights operational constraints faced by grassroots awareness initiatives and the institutional failure to treat FGM as a human rights violation, potentially fostering impunity.

## Discussion

Previous studies believed that awareness programs have played a significant role in challenging traditional beliefs and reshaping behaviors related to FGM.^58 120^ The study revealed that these programs are particularly effective in urban areas, where schools and public health campaigns provide structured opportunities for education. Awareness campaigns often focus on the physical and psychological consequences of FGM,^11^ such as its health risks and impact on mental well-being (Latha et al., 2020). These messages are tailored to dispel myths, including the belief that FGM is necessary for preserving chastity or preventing promiscuity.

The success of awareness programs in changing beliefs lies in their ability to foster critical thinking among participants,^121^ particularly young adults. Initiatives that educate women about health risks and legal prohibitions of FGM empower them to make informed decisions, aligning with the perspectives of Njue et al.^93^ and Odera et al.^99^ Furthermore, younger generations exposed to these programs often act as agents of change within their families, introducing alternative perspectives that challenge entrenched cultural norms. However, in a similar observation reported by Abdulnor,^1^ while individual beliefs may change, behavioral shifts at the community level are often slower due to persistent social pressures and deeply rooted traditions. The transfer of knowledge about the dangers of FGM to younger generations is one of the most effective strategies for reducing its prevalence.^78 90^ Schools play a pivotal role in this process,^88 91^ serving as safe spaces where adolescents can learn about FGM without fear of judgment.^134^ Awareness campaigns targeting young people often include interactive sessions, open discussions, and educational materials that emphasize the medical and legal implications of the practice.^13 14 35 72^ These efforts not only inform young adults but also create a ripple effect, as informed individuals share their knowledge with peers and family members.

Despite these successes, the study highlighted challenges in ensuring consistent knowledge transfer, particularly in rural areas. Many communities lack access to schools or structured awareness programs, leaving adolescents reliant on family elders for guidance. This challenge is particularly pronounced in low- and middle-income countries, where socio-economic factors, geographical disparities, and cultural norms create significant barriers to structured health education.^64 118^ In such cases, traditional beliefs often dominate, perpetuating the cycle of FGM. This disparity underscores the need to expand educational initiatives to underserved areas, ensuring that all young people, regardless of location, have access to accurate information about FGM. Formal education has proven to be a key factor in reducing the prevalence of FGM among girls.^17 18 22^ However, the impact of education on eliminating completely the practice may take a considerable amount of time to manifest.^78^

Geographic disparities also hinder the reach and impact of awareness programs.^91 140^ While urban areas benefit from access to schools, clinics, and media campaigns, rural communities often lack such resources. In these areas, traditional beliefs are less likely to be challenged, and awareness programs struggle to gain traction. Urban areas typically have better access to diverse media platforms, including television, the Internet, and mobile phones, which are crucial for disseminating information and challenging traditional beliefs.^65 83^ Rural areas often suffer from poor network reception and limited access to media, which hampers the effectiveness of media campaigns.^83^ Language barriers and limited access to trained facilitators, as well as logistical challenges in reaching remote locations, worsen this issue.^54 95 80^ Language barriers are a critical challenge in healthcare settings, often leading to miscommunication and reduced quality of care.^10^ Studies highlight the underutilization of interpreter services, which are sporadically used and often depend on individual healthcare practitioners’ initiative and knowledge.^42 59^

Resource constraints pose another significant barrier. Many awareness campaigns rely on funding from NGOs or government agencies,^101^ which may be inconsistent or insufficient.^57 95^ This lack of resources limits the scale and sustainability of programs,^101^ particularly in underserved areas. Additionally, the focus on short-term Greene interventions rather than long-term engagement reduces the likelihood of lasting impact.^78^ NGOs are heavily reliant on external funding, which is often short-term and insufficient for long-term sustainability.^141^ This creates a significant challenge in maintaining and expanding services to meet evolving community needs. Inadequate enforcement of anti-FGM laws further undermines the effectiveness of awareness programs.^84 122^ While legal frameworks prohibiting FGM exist,^33 36 104 122^ ^127^ their enforcement is often weak, particularly in rural areas where traditional practices are deeply ingrained. Without visible consequences for violating these laws, families may feel emboldened to continue the practice, undermining the efforts of awareness campaigns.^43 58 60^ Addressing this issue requires stronger collaboration between governments, NGOs, and community leaders to ensure that laws are effectively communicated and enforced.

Furthermore, awareness programs should incorporate a multi-generational approach that includes parents, caregivers, and community leaders alongside adolescents.^20^ ^115^ Families are integral to community structure, providing social ties and support systems that are essential for community cohesion.^117^ For effectiveness, the change must therefore begin at the family level, with parents, especially fathers, taking a more active role in their daughters’ lives.^91^ By addressing the perspectives of all stakeholders, these initiatives can create a supportive environment for rejecting FGM.^38 96^

### Study Limitation

The study was confined to participants in Ibadan, Nigeria, perhaps limiting the applicability of the results to other regions or nations with distinct cultural and socioeconomic dynamics. Given that the data were self-reported, there exists a potential for individuals to underreport or exaggerate their reactions due to social desirability biases or the sensitivity associated with FGM. Although qualitative data offer depth, the lack of accompanying quantitative data restricts the capacity to assess the statistical significance of the findings or trends.

Furthermore, due to the geographical separation between the researcher and the participants, the ethnographic research method, which emphasises the cultural viewpoints of individuals within a context, could not be employed in this study as the primary author responsible for data collection was not physically present. He was in the United Kingdom as a full-time foreign student and lacked the authorisation, time, and finances to travel to Nigeria for in-person participant recruiting or data collection. An appropriate strategy for recruiting and data collection, considering this predicament and the stringent deadline for project completion, was an online approach. Each participant was required to possess a smartphone or computer and be proficient in using MS Teams. Technologically proficient individuals constituted the cohort capable of realistically engaging in this investigation. The study participants were deemed sufficient to address the research issue.

Physically attending to recruiting people and gathering data would have enhanced the depth of the research findings through prolonged observations and the documentation of field notes alongside interviews.

## Conclusion

This study highlights the transformative potential of awareness programs and education to reduce the prevalence of FGM among young women in Ibadan. Collaborative efforts between anti-FGM advocates and educators have proven instrumental in equipping communities with knowledge to reject the practice, making education a key driver of long-term change. Studies consistently demonstrate that girls with little or no education are more likely to undergo the procedure and endorse its continuation. While these Female Genital Mutilation awareness programs have made significant strides in changing individual beliefs and behaviors, their impact is often limited by cultural resistance, geographic disparities, resource constraints, and weak legal enforcement. Incorporating anti-FGM messaging into school curricula can address this gap by fostering informed discussions in safe learning environments.

By integrating FGM awareness into both healthcare services and educational systems, these efforts ensure that anti-FGM messaging reaches multiple facets of community life, creating a comprehensive approach to prevention and stigma reduction.

Similarly, Oyo state and the federal governments must prioritize the implementation of legal frameworks, ensuring that violations are met with appropriate consequences. Collaboration with local leaders can enhance compliance, as community members are more likely to respect laws endorsed by trusted figures. Additionally, publicizing the existence and enforcement of these laws through awareness campaigns can deter families from practicing FGM.

Overall, integrating FGM awareness into broader public health and education initiatives can enhance the sustainability of these programs. For example, incorporating anti-FGM messaging into reproductive health services or school curricula ensures that information about FGM becomes a routine part of community life. This integration also helps normalize discussions about FGM, reducing the stigma associated with addressing the issue.

## Acknowledgements

We want to acknowledge and express our profound gratitude to Bukola Christianah, family, and friends for their unwavering encouragement and support at all times, and also to all the participants who took part in this study for sharing their valuable experience and perception with us.

## Contributors

The first author, S.G., was actively involved in the research from the beginning, when they wrote the study proposal and applied for ethics approval. He helped gather and analyse data and write the final study report. The second author, E.J., was also completely involved in the research process. He oversaw every step of the study from the beginning and supplied both theoretical and practical advice. Both writers read the article before it was submitted.

## Patient and public involvement

Patients and/or the public were not involved in the design, or conduct, or reporting, or dissemination plans of this research.

## Consent to Participate

A form was filled out, and consent was obtained online from all participants before the interviews, after approval from the University of Hertfordshire Health, Science, Engineering, and Technology Ethics Committee. All participants’ information provided in the results section is coded. The consent that was obtained from all of the participants was informed. The study adhered to the Declaration of Helsinki to this regard. Refer to the Methods section for further details.

## Consent for Publication

Not applicable.

## Ethics Approval

Ethical approval for the study was granted by the University of Hertfordshire Health, Science, Engineering, and Technology Ethics Committee (Protocol number: cLMS/PGT/UH/05727; dated August 12, 2024). The consent that was obtained from all of the participants was informed. The study adhered to the Declaration of Helsinki to this effect.

## Data availability

The datasets generated and analysed during the current study are not publicly available due to ethical constraints but are available from the corresponding authors on reasonable request.

## Interview guide

The interview guide used was developed for this study.

